# Depression and Complicated Grief Among Parents of Pediatric Cancer Patients in Cameroon: Implications for Global Health in Low-income Countries

**DOI:** 10.1101/2025.03.28.25324833

**Authors:** Isaac Che Ngang, Francine Kouya, Emmanuel Tetteh, Clifford Atuiri, Kaah Joel, Bienvenue Kouya, Nyinya Valery Ngalle, Vaishnavi Mamillapalle, Robert M. Chamberlain, Amr S. Soliman

## Abstract

**Background and Objectives:** Bereaved parents of pediatric cancer patients frequently experience severe grief and psychological distress, but studies on the prevalence of major depressive disorder (MDD) and complicated grief (CG) among this population in Cameroon are lacking. This study aimed to determine the prevalence of MDD and CG among bereaved parents of deceased pediatric cancer patients treated at Mbingo Baptist Hospital Cameroon, and to identify predictors of these mental health outcomes.

**Methods:** This cross-sectional study included parents of deceased pediatric cancer patients treated at Mbingo Baptist Hospital between January 2015 and January 2022. Multivariate logistic regression identified predictors of MDD and CG as adjusted odds ratios (AOR) with 95% confidence intervals (CIs).

**Results:** The prevalence of CG in this population was 86%, while 66.7% of the study subjects screened positive for MDD. Significant predictors of MDD included age [OR 1.091, p=0.018], financial hardship [OR 9.47, p=0.014], accurate knowledge of the child’s prognosis [OR 0.268, p=0.046], perceived social support [(poor social support OR 6.402, P=0.039), (moderate social support OR 8.556, p=0.045)], and coping capacity [medium resilient copers OR 7.874, p=0.027]. Predictors of CG included age [OR 1.157, p=0.032], financial hardship [OR 11.501, p=0.04], years passed since child loss [1-2 years OR 4.634, p=0.049], and coping capacity [(low resilient copers OR 14.011, p<0.01), (medium resilient copers OR 19.023, p<0.01)].

**Conclusions:** The study revealed high prevalence of MDD and CG among bereaved parents of pediatric cancer patients in Cameroon. Financial difficulty, social support, and coping capacity had substantial impact on parental mental health outcomes in this population. Personalized mental health support services into pediatric oncology care is critical for assisting bereaved families and encouraging resilience in the face of loss may improve health and wellbeing of the families. The study may have implications for global mental health in similar low-income countries.

## INTRODUCTION

Childhood cancers account for about 5% of all cancers in Africa, compared to the worldwide proportion of 1%^1^. In Cameroon, the incidence rate of childhood cancers stands at 25 cases per 1,000,000 children below 15 years annually. Children in this age group make up 42% of the Cameroon’s 27 million inhabitants, meaning each year, approximately 325 children in Cameroon are diagnosed with cancer^1^. Burkitt’s lymphomas are the most common childhood cancers in Cameroon with an incidence rate of 3.7 per 100,000 children annually. Other common childhood cancers in Cameroon include lymphomas, rhabdomyosarcomas, Wilms tumors, retinoblastomas, and Kaposi sarcomas^2^. Tragically, due to late diagnosis, limited treatment options, and financial constraints, nearly 80% of these children succumb to their illness within five years of diagnosis^2,3^. A child’s death is a devastating and unimaginable event, which usually leaves the surviving parents feeling helpless, guilty, shocked and in profound grief^3^. Bereavement grief is the term used to describe the constellation of debilitating and painful emotional, cognitive, behavioral, and somatic symptoms following the loss of a loved one^4^. It is a common, universal, physical, and psychological reaction to loss, and for most individuals typically decreases in intensity over time^5^. However, for a significant minority of individuals, the grieving process complicates into major depressive disorder (MDD), suicidal ideation^6^, or complicated grief (CG)^7^ also called prolonged grief disorder, defined as intense grief for more than 6 months.

The stress of caring for a severely ill loved one, coupled with their subsequent death predisposes bereaved parents to Major depressive disorder and Complicated Grief, and prevalence’s as high as 34% and 30% respectively have been reported in recent literature^6,7^. In Cameroon, as in most other countries in sub-Saharan Africa, parents often assume the primary caregiving role, staying with their children throughout hospital admissions, managing treatment regimens, and providing emotional support^1^. This caregiving responsibility, deeply rooted in cultural expectations, places significant psychological, social, and financial burdens on these parents, who often struggle with grief and mental health challenges long after their child’s passing^4,7^. Parents with CG suffer from increased risk of chronic physical and mental morbidities, with overall increased mortality rates^3^. One might as well say “they die from a broken heart”. Despite the relatively higher childhood cancer mortality rates in Cameroon, and the adverse health outcomes associated with parental grief, there is currently no study to the best of our knowledge aimed at measuring the prevalence of Major depressive disorder and complicated grief among this group of parents in Cameroon.

To gain some insight into this issue, we set out to determine prevalence of MDD and CG among bereaved parents of deceased pediatric cancer patients treated at Mbingo Baptist Hospital Cameroon, and to evaluate predictors of MDD and CG among the study participants.

## METHODS

### Study Design

This cross-sectional study included parents or primary caregivers of deceased pediatric cancer patients who had been treated at the Mbingo Baptist Hospital Cancer treatment center in Cameroon between January 2015 and January 2022. We obtained ethical clearance from the Cameron Baptist Convention Institutional Review Board. Eligible participants were identified by chart review, and the families were contacted by the hospital Chaplain who obtained verbal informed consent to the study.

### Study site

Mbingo Baptist Hospital is a 310-bed referral facility in the Northwest region of Cameroon, with 10 beds dedicated for in-patient treatment of pediatric cancer cases. Mbingo Baptist Hospital is one of the only two cancer treatment centers serving the Northwest region of the country.

### Subjects

We recruited parents or primary caregivers of deceased pediatric cancer patients who were at least 6 months post-loss, aged 18 or older, not on treatment for any mental health condition, and who gave their consent. We defined a childhood cancer patient as someone diagnosed with cancer by a physician in a health facility, and they were younger than 15 years at the time of diagnosis.

Primary caregivers included had been closely involved with the care and welfare of the child. For each child, only one parent or primary caregiver was recruited. Our sampling frame contained 105 eligible participants. To attain a 95% statistical power, we needed a minimum sample size of 83 participants. We accounted for a non-response rate of 10%, added 9 participants and set the new minimum sample size to 92 participants.

### Outcome variables

The outcome variables were major depressive disorder (MDD) and complicated grief (CG). We measured MDD using the Patient Health Questionnaire-9 (PHQ-9)^8^. This is a 9-item tool based on the 9 diagnostic criteria for MDD in the DSM-IV. Responses are scored on a Likert scale from 0 (not at all) to 3 (nearly every day). Final scores of 5, 10, 15, and 20 represent cutoffs for mild, moderate, moderately severe, and severe depression respectively.

We measured CG using the Brief Grief Questionnaire^9^. This is a standardized 5-item instrument used to screen for CG among bereaved individuals. The responses for each item are graded on a Likert scale from 0 (not at all) to 2 (a lot). A final score of 4 or more suggests that an individual may be suffering from CG and should prompt a referral to a grief specialist for further evaluation.

### Independent variables

We identified several variables from current literature which had been predictive of MDD and/or CG among study participants in settings like ours ^10-15^.

Perceived social support; this was measured using the Oslo Social Support Scale (OSSS-3)^16^. This is a 3-item instrument that asks for number of close confidants, level of concern from other people and relationship with neighbors.

Resilience: this was measured using the brief resilience coping scale^17^, a 4-item instrument with 5-point scale responses and a total score ranging from 4-20. Participants who score between 4-13 are considered low resilient copers, those who score between 14-16 are considered medium resilient copers and those who score between 17-20 are high resilient copers.

Caregiver burden: this was measured using the short version of the Burden Scale for Family Caregivers (BCFC-s)^18^. This is a standardized highly reliable tool (Chronbach’s alpha = 0.92), with translations in 20 European languages indicating that it could be applied to a variety of settings. The questionnaire includes 10 questions with each question’s responses scored from 0-3. Scores were reported on a continuous scale with a possible range from 0-30.

Financial hardship: participants responded to the question ‘would you say you are experiencing financial hardship?’ with a ‘yes/maybe’ or a ‘no’.

Extent of suffering before death: participants described their perception of the extent of their child’s suffering before death as either minimal, moderate or extreme.

Accurate knowledge of child’s prognosis; this was measured by asking participants if they knew their child’s prognosis, and if so whether the taught the disease was curable, could reoccur, or that the child was probably going to die. Accurate knowledge of child’s prognosis was defined as participants choosing the latter option.

### Data collection

This was a quantitative study that involved the use of paper-based questionnaires. Ethical clearance was obtained from the Cameroon Baptist Convention’s Institutional Review Board on April 13, 2023. Chart review and participant recruitment began on May 10, 2023 and continued till July 12 2023. The questionnaire was tested for clarity and appropriateness in a pilot study involving 5 participants. Eligible participants were identified by reviewing charts of deceased children who had received treatment at the center between January 2015 and January 2022. The chaplain and palliative care nurse reached out to eligible participants by phone calls to present the study and obtained verbal consents for home visits. Verbally consented participants were administered the questionnaires in their homes by the chaplain and the palliative care nurse. Data collected was transferred into MS Excel, exported, and analyzed in SPSS version 28.

### Data analysis

We developed a multivariable binary logistic regression model in SPSS to identify Individual predictors. The dependent variables were Complicated Grief and Major Depressive Disorder, both of which were measured as binary variables. The model included all 7 predictor variables initially selected. The regression parameter for each predictor was reported as adjusted odds ratio (AOR) with a 95% level of confidence (CI = 95%).

## RESULTS

### Participant characteristic

92 participants completed the questionnaire. The mean age of the deceased at time of diagnosis was 10.7 years (S.D = 2.2). Proportion of unemployed participants was 56.5% (n=52), 65.6% (n=61) had some primary education and the overwhelming majority (90.1%, n=82) were Christians. Most participants had lost 2 children to all-causes (74.2%, n=66), and the responses to ‘number of living children’ were evenly distributed across 1-2 children (26.1%), 3-4 children (29.3%), and 5 or more children (34.8%) respectively. Most participants reported that the loss of their child to childhood cancer had occurred within the prior 2 years (Table 1).

**Table 1:** Sociodemographic characteristics of participants.

|  | **n** | **%** |
| --- | --- | --- |
| **Participant's age, mean (SD)** | 43 | 12 |
| **Participant's employment status** |  |  |
| Full time | 11 | 12.0 |
| Part time | 29 | 31.5 |
| Unemployed/retired | 52 | 56.5 |
| **Participant's educational level** |  |  |
| No formal education | 15 | 16.1 |
| Some primary education | 61 | 65.6 |
| Some secondary education | 14 | 15.1 |
| Some university education | 2 | 3.2 |
| **Financial hardship** |  |  |
| Yes | 74 | 81.3 |
| No | 17 | 18.7 |
| **Participant's religion** |  |  |
| Christian | 82 | 90.1 |
| Muslim | 8 | 8.8 |
| Traditional worship | 2 | 1.1 |
| Atheism | 0 | 0 |
| **Number of children lost** |  |  |
| 1 child | 11 | 12.4 |
| 2 children | 67 | 74.2 |
| 3 children | 10 | 10.9 |
| 4 or more children | 4 | 4.3 |
| **Living children** |  |  |
| None | 9 | 9.8 |
| 1 – 2 children | 24 | 26.1 |
| 3 – 4 children | 27 | 29.3 |
| 5 or more children | 32 | 34.8 |
| **Participant's marital status** |  |  |
| Single/divorced | 28 | 30.8 |
| Married | 52 | 57.1 |
| Widowed | 12 | 12.1 |
| **Deceased child's sex** |  |  |
| Male | 46 | 50 |
| Female | 46 | 50 |
| **Child's age at time of diagnosis** |  |  |
| Less than 1 year | 3 | 3.3 |
| 1 – 5 years | 23 | 25.0 |
| 6 – 10 years | 21 | 22.8 |
| 11 – 15 years | 45 | 48.9 |
| **Years passed since child loss** |  |  |
| Less than 1 year | 22 | 23.7 |
| 1 – 2 years | 25 | 28.0 |
| 3 – 4 years | 22 | 23.7 |
| 5 or more years | 23 | 24.7 |

### Prevalence of Major Depressive Disorder and Complicated Grief

Among study participants, 86% screened positive for CG (Figure 2). 11.8% were minimally depressed, and 18.3 were mildly depressed. 36.6% and 30.1% of participants screened positive for moderate and severe depression respectively (Figure 3). When the depression variable was dichotomized into MDD positive or not, 66.7% (n=62) of participants screened positive for MDD (Figure 2).

**Fig 1.**
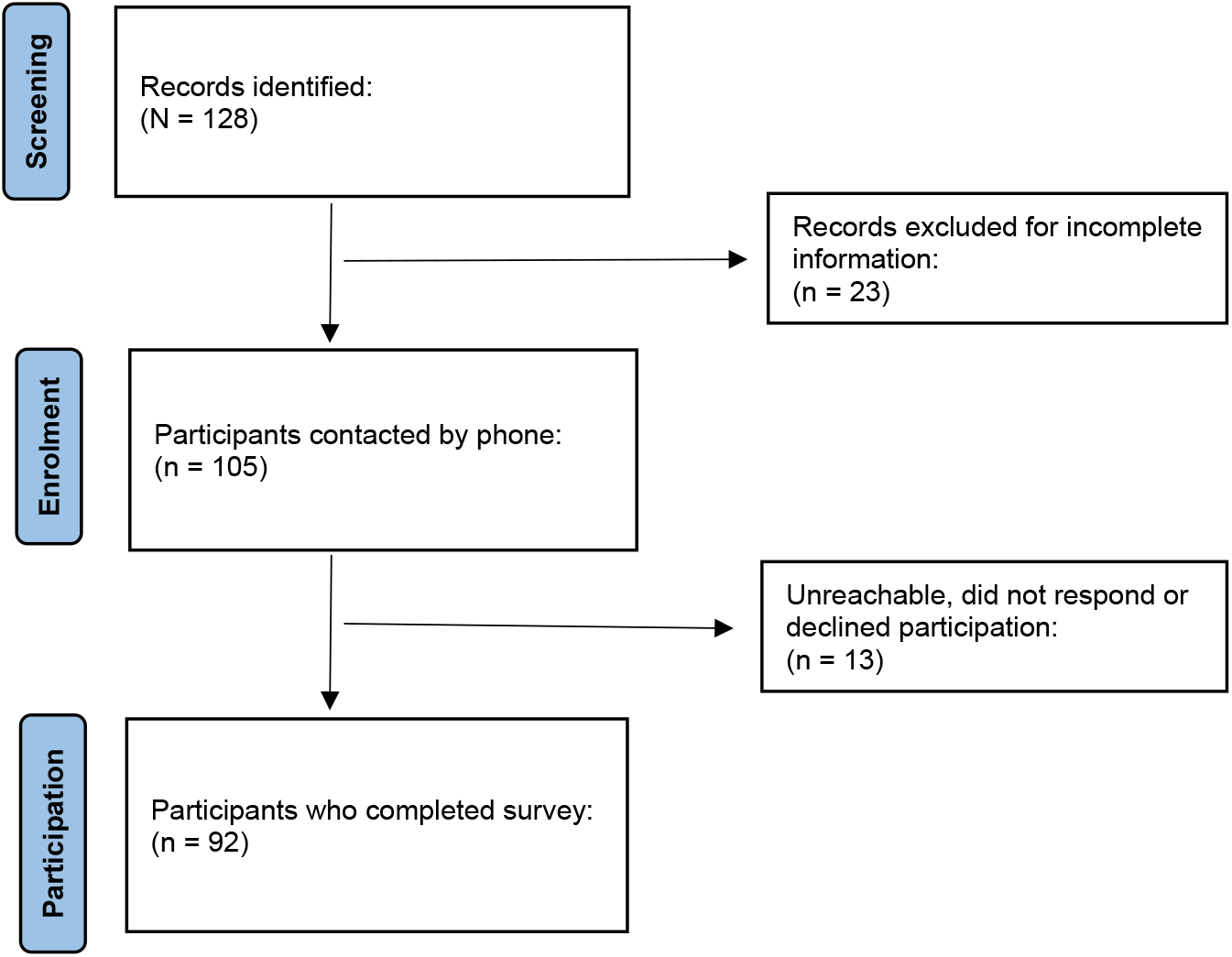
Flow chart of participant screening and enrollment.

**Fig 2.**
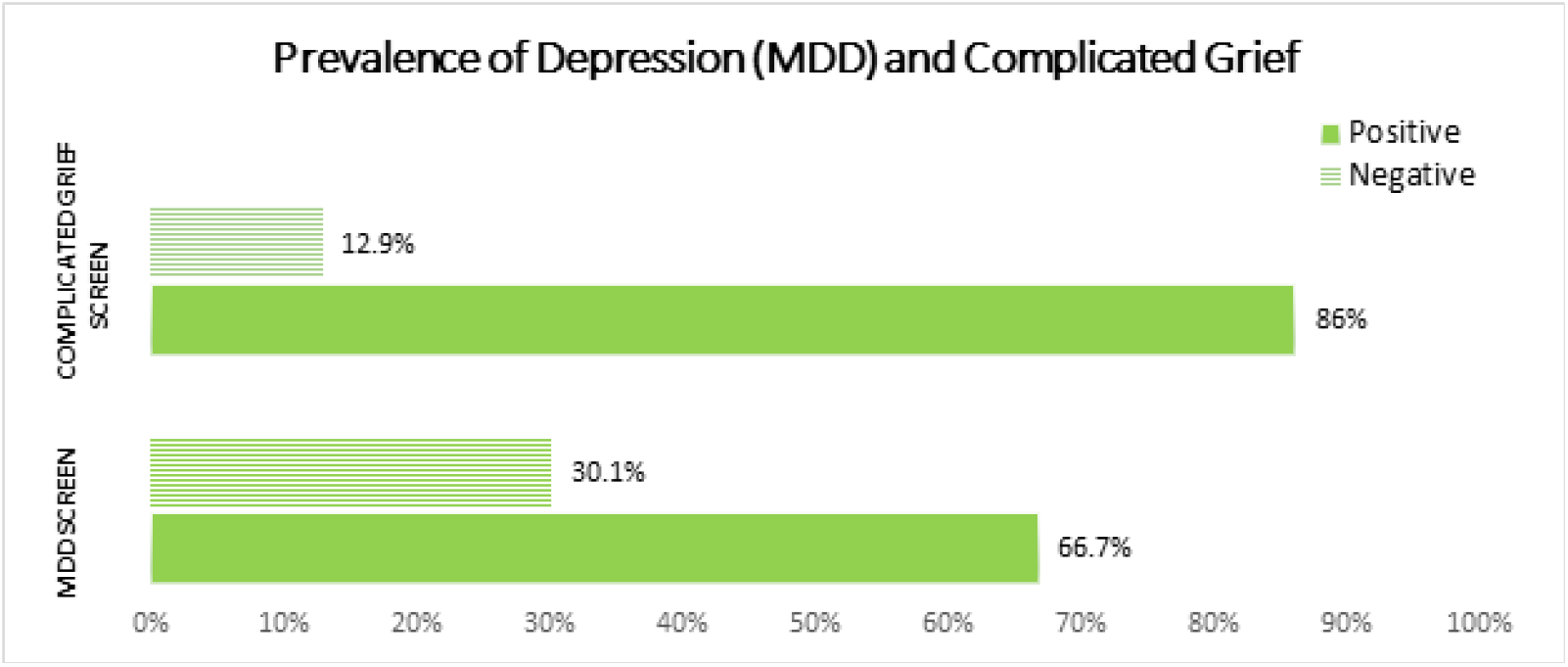
Prevalence of Major Depressive Disorder (MDD) and Complicated Grief.

**Figure 3:**
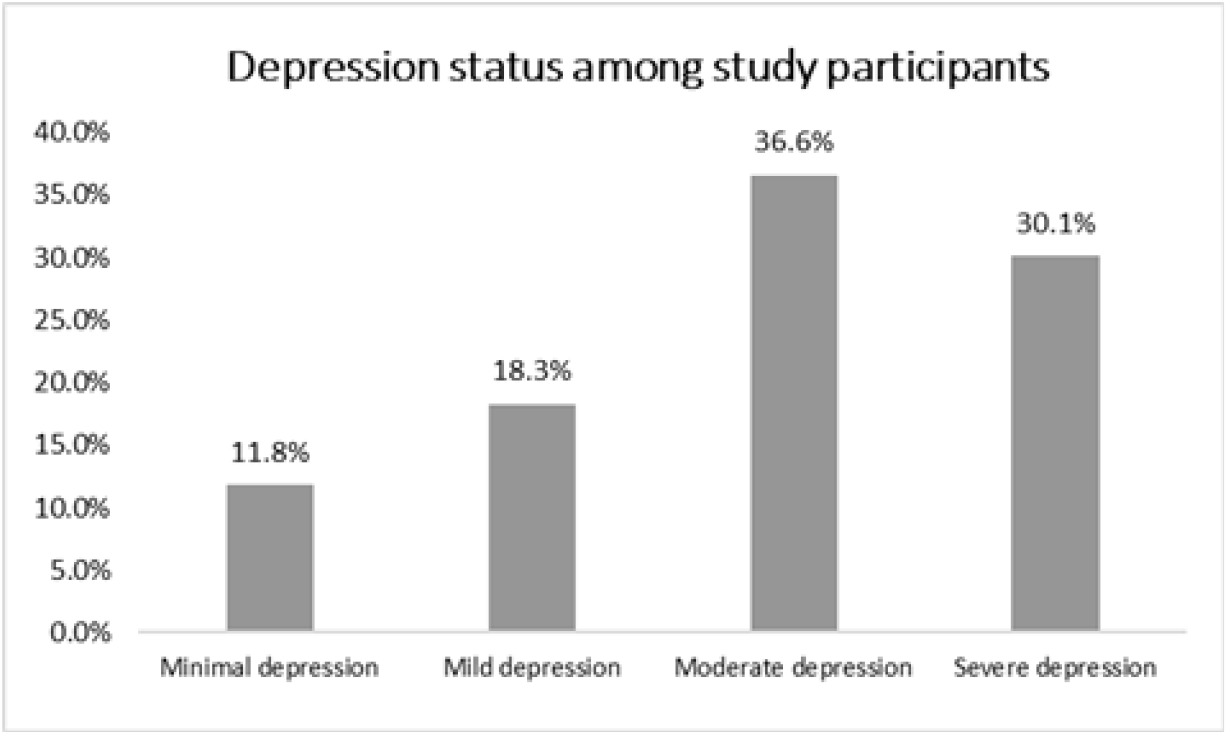
Depression types among study participants.

### Predictors for Major Depressive Disorder

Among study participants, age (aOR = 1.091, CI: 1.015-1.174, p=0.018), financial hardship (aOR 9.47, CI: 1.584 – 56.629, p=0.014), accurate knowledge of child’s prognosis at diagnosis (aOR 0.268, CI: 0.073 – 0.977), perceived social support and coping capacity were significant predictors of MDD. Participants who reported moderate and poor social supports were about 8 times (aOR 8.556 CI: 1.049-69.809) and 6 times (aOR 6.402 CI: 1.095 – 37.432) respectively more like to screen positive for MDD compared to those who reported strong social support. Medium-resilient and low-resilient copers were about 8 times (aOR 7.874 CI: 1.385 – 23.728, P = 0.027) and (aOR 8.401 CI: 0.854-82.686, P=0.068) respectively more likely to screen positive for MDD compared to high copers, although the latter was not statistically significant. Contrary to findings in current literature, years passed sine child loss, perception of extent of suffering before death and the burden of caregiving were not predictive of MDD among study participants (Table 2).

**Table 2:** Predictors of Major Depressive Disorder and Complicated Grief.

| Independent variable | Major Depressive Disorder |  |  | Complicated grief |  |  |
| --- | --- | --- | --- | --- | --- | --- |
|  | AOR | 95% CI | p value | AOR | 95% CI | p value |
| **Age** | 1.091 | 1.015 – 1.174 | **0.018** | 1.157 | 1.012 – 1.322 | **0.032** |
| **Financial hardship** |  |  |  |  |  |  |
| Yes | 9.47 | 1.584 – 56.629 | **0.014** | 11.501 | 1.115 – 118.664 | **0.04** |
| No | Ref | Ref |  | Ref | Ref |  |
| **Years passed since child loss** |  |  |  |  |  |  |
| 1 year | 2.443 | 0.424 – 14.086 | 0.318 | 5.860 | 0.191 – 180.249 | 0.312 |
| 1 – 2 years | 3.490 | 0.585 – 20.812 | 0.170 | 4.634 | 1.012 – 210.191 | **0.049** |
| 3 – 4 years | 1.920 | 0.363 – 10.171 | 0.443 | 5.173 | 0.520 – 51.477 | 0.161 |
| 5 or more years | Ref | Ref |  | Ref | Ref |  |
| **Accurate knowledge of child's prognosis at diagnosis** |  |  |  |  |  |  |
| Yes | 0.268 | 0.073 – 0.977 | **0.046** | 0.195 | 0.016 – 2.441 | 0.205 |
| No | Ref | Ref |  | Ref | Ref |  |
| **Perception of extent of suffering before death** |  |  |  |  |  |  |
| Minimal | 0.039 | 0.001 – 1.217 | 0.065 | 2.285 | 0.001 – 1142.402 | 0.849 |
| Moderate | 0.514 | 0.085 – 3.099 | 0.468 | 0.761 | 0.037 – 15.523 | 0.859 |
| Extreme | Ref | Ref |  | Ref | Ref |  |
| **Perceived social support** |  |  |  |  |  |  |
| Poor social support | 6.402 | 1.095 – 37.432 | **0.039** | 1.896 | 0.199 – 17.584 | 0.585 |
| Moderate social support | 8.556 | 1.049 – 69.809 | **0.045** | 2.163 | 0.117 – 40.032 | 0.604 |
| Strong social support | Ref | Ref |  | Ref | Ref |  |
| **Coping capacity** |  |  |  |  |  |  |
| Low resilient copers | 8.401 | 0.854 – 82.686 | 0.068 | 14.011 | 1.136 – 156.867 | **<0.01** |
| Medium resilient copers | 7.874 | 1.385 – 23.728 | **0.027** | 19.023 | 2.537 - 109.001 | **<0.01** |
| High resilient copers | Ref | Ref |  | Ref | Ref |  |
| **Caregiver burden score** | 1.049 | 0.983 – 1.119 | 0.149 | 1.105 | 0.981 – 1.245 | 0.1 |

### Predictors for Complicated Grief

Age (aOR 1.157 CI: 1.084 - 1.230, p = 0.032), financial hardship (aOR 11.501, CI: 1.115 – 118.664, p = 0.046), years passed since child loss and coping capacity were significant predictors of screening positive for CG among study participants. Low resilient copers times (aOR 0.014 CI: 0.001 – 0.156, P<0.01) and medium resilient copers (aOR 0.019 CI: 0.002 – 0.019, p<0.01 were less likely to screen positive for CG compared to high resilient copers. Participants whose children had passed in the past 1-2 years were about 5 times (aOR 4.634 CI: 1.1012 – 210.191, P=0.049) more likely to screen positive for CG compared to those whose child had passed 5 or more years ago. Participants at 1-year and those at 3-4 years post loss did not have significantly higher risks for screening positive for CG (Table 2).

## DISCUSSION

Each year, at least 325 children in Cameroon are diagnosed with pediatric cancers, the most frequent of which are Burkitt lymphomas. Unfortunately, four out of every five of these youngsters will not live more than five years after their diagnosis. The traumatic impact of losing a child to cancer, along with the lack of proper support systems and resources, places enormous emotional and psychological loads on the surviving parents, predisposing them to MDD and CG. This is the first study to investigate the prevalence of MDD and CG among bereaved parents of pediatric cancer patients in Cameroon, as well as some common predictors of these outcomes.

### Prevalence of Major Depressive Disorder and Complicated Grief

Our study’s findings reveal a disturbingly high prevalence of MDD and CG respectively among bereaved parents. Approximately two-thirds of the grieving parents screened positive for MDD, while an overwhelming 86% had symptoms consistent with CG. These figures reaffirm the significant emotional suffering faced by parents after losing a child to cancer. Our study’s prevalence of MDD and CG substantially exceeds the national depression rate of 8.4% in Cameroon reported by Fodjo et al^19^ in 2021, underscoring the need for tailored interventions to support bereaved parents in Cameroon.

Other studies that examined depression and cg rates among bereaved family members and spouses found values ranging from 20-40% for CG^7,20-21^ and 26-34% for MDD^6,22-23^. Possible explanations for these significant differences include stronger attachments between parents and their children, the absence of any form of post-bereavement service among our participants, a practice that is very common in the West, where most reported studies were conducted, and our use of screening rather than diagnostic tools in our study. Despite the disparities in raw numbers, these studies convey the same story: mourning increases the probability of negative mental consequences after loss, emphasizing the need for focused therapies for bereaved parents.

### Predictors of Major Depressive Disorder and Complicated Grief

Several predictors emerged as significant contributors to the development of MDD and CG among the study participants. Financial hardship, accurate knowledge of the child’s prognosis at diagnosis, perceived social support, and coping capacity were among the key factors influencing parental mental health outcomes.

Financial stress appeared as a major predictor of both MDD and CG, emphasizing the link between socioeconomic issues and psychological well-being. Parents experiencing financial challenges may face increased stress and a sense of helplessness, increasing their susceptibility to mental health illnesses. Addressing financial barriers to getting healthcare and giving financial assistance to impacted families may ease some of the burdens that contribute to depressive symptoms and complicated grief disorder. These findings mirror those of Wen at al.^11^ in a 2022 study which showed that personal vulnerability (measured as financial hardship) among bereaved caregivers predisposed them to more distressing depressive symptom trajectories.

Accurate awareness of the child’s prognosis at the time of diagnosis was linked to a lower risk of MDD, highlighting the significance of open and honest communication between healthcare personnel and families. Parents who are well-informed about their child’s prognosis may feel more prepared and accepting, thus reducing psychological suffering caused by uncertainty and ambiguity around the outcome of their child’s condition. Our finding differed from those of Wen et al.^11^, who found no relationship between accurate knowledge of child’s prognosis and post-loss depression. More than eighty percent of our participants were religious. As a result, it is probable that they received signals of hope and recovery for their children during the treatment phase, which exacerbated their frustration when the child died.

Perceived social support emerged as a significant protective factor against MDD, with participants reporting strong social support being less likely to screen positive for depression. Previous literature has emphasized that social support helps in the grieving process^25^, and a lack of social support is a risk factor for negative bereavement outcomes^26^. According to Tompson et al.^27^, bereaved parents and siblings rely on family for social support from 6 to 19 months after losing a child to cancer. Pediatric oncology units play a vital role in assisting families during the palliative and grieving stages. A bereavement program should provide formal support services, communication from medical staff throughout palliation and after the child’s death, and opportunities to connect with other cancer-bereaved families for support^28^. Similarly, Nolbris and Hellström^29^ reported that friends are a vital source of social support for the bereaved.

Coping capacity, as evaluated by resilience, was also an important factor in predicting mental health outcomes. High resilient copers were less likely to test positive for CG than medium and low resilient copers. Building resilience through tailored interventions and psychotherapy approaches may help parents navigate the grief process more successfully and adaptively. These findings mirror those of Rasouli et al.^15^ in their 2022 study in which they found that higher resilience (measured by personal competence scores) was associated with lower rates of CG among bereaved siblings.

### Implications for practice and policy

The study’s findings have significant implications for clinical practice, policy creation, and resource allocation in pediatric cancer care in Cameroon and other similar countries. To begin, there is an urgent need to incorporate mental health support services into pediatric cancer care to meet the psychological needs of grieving parents. This could include educating healthcare providers about psychosocial care, developing grief support groups, and providing access to counseling and therapy services.

Furthermore, initiatives to improve communication and information exchange between healthcare practitioners and families can help to make better informed decisions and boost psychological readiness for end-of-life care. Investing in comprehensive palliative care services that stress holistic support for patients and their families, such as psychosocial and spiritual care, is critical to enhancing the quality of life for children with cancer and their caregivers.

On a policy level, there is a need to increase awareness of the psychosocial aspects of pediatric cancer care, as well as the necessity of mental health support for impacted families. Advocacy activities to raise mental health awareness, decrease stigma, and devote resources for psychosocial support services are crucial to meeting the unmet needs of bereaved parents and encouraging resilience in the face of loss.

### Study limitations

First, the participants were limited to parents who had lost a child at the Mbingo Baptist Hospital facility; therefore, the findings may not represent the entire population. Second, because of the nature of the study design, we cannot rule out recall bias. Third, the subjects who refused participation may have been suffering from severe grief or depression, although the response rate in the present study was 87.6%, which is a relatively high rate compared with previous studies^30^. Finally, assessing CG and MDD, we used the BGQ and the PHQ-9, which are both screening tools and therefore may have over-estimated the true prevalence of MDD and CG among our study participants.

## CONCLUSION

Finally, this study sheds light on the tremendous emotional impact of children cancer on parents and caregivers in Cameroon. Identifying predictors of MDD and CG in bereaved parents gives useful insights for developing tailored therapies to help families cope with the death of a child to cancer. Addressing affected families’ psychosocial needs must be a top focus in pediatric oncology care, with coordinated efforts required to include mental health support services into existing healthcare systems and regulations.

## Data Availability

Data will be made available upon reasonable request to the corresponding author.

## Acknowledgement

Isaac Che Ngang was supported by the Cancer Epidemiology Education in Special Populations (CEESP) Program through funding from the National Institute grant # R25CA112383.

## Appendix 1. Survey Instrument

## Appendix 2. Codebook

## Appendix 3. Cameroon Baptist Convention IRB Approval letter

## References

1. Afungchwi GM, Kruger M, Wharin PD, Bardin R, Kouya FN, Hesseling PB. The Evolution of a Hospital-Based Cancer Registry in Northwest Cameroon from 2004 to 2015. J Trop Pediatr. 2021;67(3). doi:10.1093/tropej/fmaa038

2. Afungchwi GM, Kruger M, Kouya F, et al. Two decades of childhood cancer care in Cameroon: 2000–2020. Pediatr Blood Cancer. 2021;68(7). doi:10.1002/pbc.28997

3. Snaman JM, Kaye EC, Torres C, Gibson D v., Baker JN. Helping parents live with the hole in their heart: The role of health care providers and institutions in the bereaved parents’ grief journeys. Cancer. 2016;122(17):2757–2765. doi:10.1002/cncr.30087

4. Wen FH, Prigerson HG, Chou WC, et al. Prolonged grief disorder and depression are distinguishable syndromes: A latent transition analysis for bereaved family caregivers of cancer patients. Psychooncology. 2022;31(7):1144–1151. doi:10.1002/pon.5902

5. Lundorff M, Holmgren H, Zachariae R, Farver-Vestergaard I, O’Connor M. Prevalence of prolonged grief disorder in adult bereavement: A systematic review and meta-analysis. J Affect Disord. 2017;212:138–149. doi:10.1016/j.jad.2017.01.030

6. Aoyama M, Miyashita M, Masukawa K, et al. Factors related to suicidal ideation among bereaved family members of patients with cancer: Results from a nationwide bereavement survey in Japan. J Affect Disord. 2022;316:91–98. doi:10.1016/j.jad.2022.08.019

7. Guldin MB, Vedsted P, Zachariae R, Olesen F, Jensen AB. Complicated grief and need for professional support in family caregivers of cancer patients in palliative care: A longitudinal cohort study. Supportive Care in Cancer. 2012;20(8):1679–1685. doi:10.1007/s00520-011-1260-3

8. Kroenke K, Spitzer RL, Williams JB. The PHQ-9: validity of a brief depression severity measure. J Gen Intern Med. 2001 Sep;16(9):606–13. Doi:10.1046/j.1525-1497.2001.016009606.x. PMID: 11556941; PMCID: PMC1495268

9. Ito M, Nakajima S, Fujisawa D, Miyashita M, Kim Y, Shear MK, Ghesquiere A, Wall MM. Brief measure for screening complicated grief: reliability and discriminant validity. PloS One. 2012;7(2):e31209. doi: 10.1371/journal.pone.0031209. Epub 2012 Feb 14. PMID: 22348057; PMCID: PMC3279351.

10. Große J, Treml J, Kersting A. Impact of caregiver burden on mental health in bereaved caregivers of cancer patients: A systematic review. Psychooncology. 2018 Mar;27(3):757–767. doi: 10.1002/pon.4529. Epub 2017 Sep 7. PMID: 28805954.

11. Wen FH, Chou WC, Prigerson HG, Shen WC, Hsu MH, Tang ST. Predictors of Family Caregivers’ Depressive- and Prolonged-Grief-Disorder-Symptom Trajectories. J Pain Symptom Manage. 2022 Apr;63(4):476-484.e1. doi: 10.1016/j.jpainsymman.2021.12.025. Epub 2021 Dec 29. PMID: 34971750.

12. Wen FH, Chou WC, Hou MM, Su PJ, Shen WC, Chen JS, Chang WC, Hsu MH, Tang ST. Associations of death-preparedness states with bereavement outcomes for family caregivers of terminally ill cancer patients. Psychooncology. 2022 Mar;31(3):450–459. doi: 10.1002/pon.5827. Epub 2021 Sep 29. PMID: 34549848.

13. Wen FH, Chou WC, Hou MM, Su PJ, Shen WC, Chen JS, Chang WC, Tang ST. Depressive-Symptom Trajectories From End-of-Life Caregiving Through the First 2 Bereavement Years for Family Caregivers of Advanced Cancer Patients. J Pain Symptom Manage. 2021 Oct;62(4):699–708. doi: 10.1016/j.jpainsymman.2021.03.018. Epub 2021 Mar 29. PMID: 33794300.

14. Hayashi Y, Sato K, Ogawa M, Taguchi Y, Wakayama H, Nishioka A, Nakamura C, Murota K, Sugimura A, Ando S. Association Among End-Of-Life Discussions, Cancer Patients’ Quality of Life at End of Life, and Bereaved Families’ Mental Health. Am J Hosp Palliat Care. 2022 Sep;39(9):1071–1081. doi: 10.1177/10499091211061713. Epub 2021 Dec 23. PMID: 34939852.

15. Omid Rasouli, Unni Karin Moksnes, Trude Reinfell, Odin Hjemdal, Mary-Elizabeth Bradley Eilertse. BMC Palliative Care (2022) 21:93 10.1186/s12904-022-00978-5.

16. Kocalevent RD, Berg L, Beutel ME, Hinz A, Zenger M, Härter M, Nater U, Brähler E. Social support in the general population: standardization of the Oslo social support scale (OSSS-3). BMC Psychol. 2018 Jul 17;6(1):31. doi: 10.1186/s40359-018-0249-9. PMID: 30016997; PMCID: PMC6050647.

17. Fung SF. Validity of the Brief Resilience Scale and Brief Resilient Coping Scale in a Chinese Sample. Int J Environ Res Public Health. 2020 Feb 16;17(4):1265. doi: 10.3390/ijerph17041265. PMID: 32079115; PMCID: PMC7068432.

18. Graessel E, Berth H, Lichte T, Grau H. Subjective caregiver burden: validity of the 10-item short version of the Burden Scale for Family Caregivers BSFC-s. BMC Geriatr. 2014 Feb 20;14:23. doi: 10.1186/1471-2318-14-23. PMID: 24555474; PMCID: PMC3942019.

19. Siewe Fodjo JN, Ngarka L, Njamnshi WY, Nfor LN, Mengnjo MK, Mendo EL, Angwafor SA, Atchou Basseguin JG, Nkouonlack C, Njit EN, Ahidjo N, Chokote ES, Dema F, Fonsah JY, Tatah GY, Palmer N, Seke Etet PF, Palmer D, Nsagha DS, Etya’ale DE, Perrig S, Sztajzel R, Annoni JM, Zoung-Kanyi Bissek AC, Leke RGF, Abena Ondoa Obama MT, Nkengasong JN, Colebunders R, Njamnshi AK. Fear and depression during the COVID-19 outbreak in Cameroon: a nation-wide observational study. BMC Psychiatry. 2021 Jul 15;21(1):356. doi: 10.1186/s12888-021-03323-x. PMID: 34266400; PMCID: PMC8280628.

20. Chiu YW, Huang CT, Yin SM, Huang YC, Chien CH, Chuang HY (2010) Determinants of complicated grief in caregivers who cared for terminal cancer patients. Support Care Cancer 18(10):1321–1327.

21. Wiese CHR, Morgenthal HC, Bartels UE, Vossen-Wellmann A, Graf BM, Hanekop GG (2010) Post-mortal bereavement of family caregivers in Germany: a prospective interview-based investigation. Wien Klin Wochenschr 122(13–14):384–389.

23. Rebollo P, Alonso J, Ramon I, Vilagut G, Santed R, Pujol R (2005) Health-related quality of life during the bereavement period of caregivers of a deceased elderly person. Qual Life Res 14(2):501–509.

24. Cherlin EJ, Barry CL, Prigerson HG, Green DS, Johnson-Hurzeler R, Kasl SV et al (2007) Bereavement services for family caregivers: how often used, why, and why not. J Palliat Med 10 (1):148–158.

25. Kreicbergs UC, Lannen P, Onelov E, Wolfe J. Parental grief after losing a child to cancer: impact of professional and social support on long-term outcomes. J Clin Oncol. 2007;25(22):3307–12.

26. Stroebe M, Stroebe W, Schut H. Gender diferences in adjustment to bereavement: an empirical and theoretical review. Rev Gen Psychol. 2001;5(1):62–83.

27. Thompson AL, Miller KS, Barrera M, Davies B, Foster TL, Gilmer MJ, Hogan N, Vannatta K, Gerhardt CA. A qualitative study of advice from bereaved parents and siblings. J Soc Work End Life Palliat Care. 2011;7(2–3):153–72.

28. deCinque N, Monterosso L, Dadd G, Sidhu R, Macpherson R, Aoun S. Bereavement support for families following the death of a child from cancer: experience of bereaved parents. J Psychosoc Oncol.

29. Nolbris M, Hellstrom AL. Siblings’ needs and issues when a brother or sister dies of cancer. J Pediatr Oncol Nurs. 2005;22(4):227–33. 2006;24(2):65–83.

30. Harrop, E.J., Morgan, F., Longo, M., Semedo, L., Fitzgibbon, J., Pickett, S., Scott, H.M., Seddon, K., Sivell, S., Nelson, A., & Byrne, A. (2020). The impacts and effectiveness of support for people bereaved through advanced illness: A systematic review and thematic synthesis. Palliative Medicine, 34, 871 – 888.

